# Comparison of the diagnostic accuracy among GPT-4 based ChatGPT, GPT-4V based ChatGPT, and radiologists in musculoskeletal radiology

**DOI:** 10.1101/2023.12.07.23299707

**Authors:** Daisuke Horiuchi, Hiroyuki Tatekawa, Tatsushi Oura, Taro Shimono, Shannon L Walston, Hirotaka Takita, Shu Matsushita, Yasuhito Mitsuyama, Yukio Miki, Daiju Ueda

## Abstract

**Objective:** To compare the diagnostic accuracy of Generative Pre-trained Transformer (GPT)-4 based ChatGPT, GPT-4 with vision (GPT-4V) based ChatGPT, and radiologists in musculoskeletal radiology.

**Materials and Methods:** We included 106 “Test Yourself” cases from *Skeletal Radiology* between January 2014 and September 2023. We input the medical history and imaging findings into GPT-4 based ChatGPT and the medical history and images into GPT-4V based ChatGPT, then both generated a diagnosis for each case. Two radiologists (a radiology resident and a board-certified radiologist) independently provided diagnoses for all cases. The diagnostic accuracy rates were determined based on the published ground truth. Chi-square tests were performed to compare the diagnostic accuracy of GPT-4 based ChatGPT, GPT-4V based ChatGPT, and radiologists.

**Results:** GPT-4 based ChatGPT significantly outperformed GPT-4V based ChatGPT (*p* < 0.001) with accuracy rates of 43% (46/106) and 8% (9/106), respectively. The radiology resident and the board-certified radiologist achieved accuracy rates of 41% (43/106) and 53% (56/106). The diagnostic accuracy of GPT-4 based ChatGPT was comparable to that of the radiology resident but was lower than that of the board-certified radiologist, although the differences were not significant (*p* = 0.78 and 0.22, respectively). The diagnostic accuracy of GPT-4V based ChatGPT was significantly lower than those of both radiologists (*p* < 0.001 and < 0.001, respectively).

**Conclusion:** GPT-4 based ChatGPT demonstrated significantly higher diagnostic accuracy than GPT-4V based ChatGPT. While GPT-4 based ChatGPT’s diagnostic performance was comparable to radiology residents, it did not reach the performance level of board-certified radiologists in musculoskeletal radiology.

## Introduction

Chat Generative Pre-trained Transformer (ChatGPT) is a novel language model based on GPT-4 architecture, which demonstrates an impressive capability for understanding and generating natural responses on various topics [1–3]. Experts in various industries have been exploring the potential applications of ChatGPT and considering how its integration could improve efficiency and decision-making processes [4]. Furthermore, the recent GPT-4 with vision (GPT-4V) enables the analysis of image inputs and offers the possibility of expanding the impact of large language models [5]. Given the potential impact of ChatGPT in the medical field, healthcare professionals need to understand its performance, strengths, and limitations for optimal utilization.

Artificial intelligence has demonstrated notable benefits in the field of radiology [6, 7], and it also holds promise for improving diagnostic accuracy and patient outcomes in musculoskeletal radiology [8, 9]. ChatGPT has the potential to be a valuable tool in improving diagnostic accuracy and patient outcomes, and there have been some initial applications of ChatGPT in radiology [10–15]. GPT-3.5 based ChatGPT nearly passed a text-based radiology examination without any specific radiology training, and then GPT-4 based ChatGPT passed the examination, [16, 17]. In musculoskeletal radiology, there has been only one study of ChatGPT, which focused on generating research articles [18].

Previous studies have evaluated the diagnostic performance of GPT-4 based ChatGPT from the patient’s medical history and imaging findings in the field of radiology [14, 15]. However, it remains unclear whether ChatGPT’s diagnostic accuracy is higher when using the images themselves or the descriptions of imaging findings. Additionally, the comparison of diagnostic performance among GPT-4 based ChatGPT, GPT-4V based ChatGPT, and radiologists has not been investigated. Current data is insufficient to determine whether the integration of ChatGPT into musculoskeletal radiology practice has the potential to improve diagnostic accuracy and reduce diagnostic errors. The journal *Skeletal Radiology* presents diagnostic cases as “Test Yourself” to allow readers to assess their diagnostic skills. These diagnostic cases offer a means to evaluate the diagnostic performance of ChatGPT in musculoskeletal radiology and obtain insights into its potential as a diagnostic tool.

This study aimed to compare the diagnostic accuracy among GPT-4 based ChatGPT, GPT-4V based ChatGPT, and radiologists in musculoskeletal radiology using the “Test Yourself” cases published in *Skeletal Radiology*.

## Materials and Methods

### Study design

This study was approved by the institutional review board of our institution, and informed consent was not required since this study utilized only published cases. We input the patient’s medical history and descriptions of imaging findings associated with each case into GPT-4 based ChatGPT, and input the patient’s medical history and images themselves associated with each case into GPT-4V based ChatGPT. Each ChatGPT generated the differential and final diagnoses, and we estimated the diagnostic accuracy rate of the outputs. Additionally, radiologists independently reviewed all the cases based on the patient’s medical history and images, and their diagnostic accuracy rates were evaluated. We then compared the diagnostic accuracy rates for the final diagnosis and differential diagnoses among GPT-4 based ChatGPT, GPT-4V based ChatGPT, and radiologists. This study was designed according to the Standards for Reporting Diagnostic Accuracy Studies statement [19].

### Data collection

The journal *Skeletal Radiology* publishes diagnostic cases in the “Test Yourself” section. We collected 128 consecutive “Test Yourself” cases from January 2014 (volume 43, issue 1) to September 2023 (volume 52, issue 9). We excluded 22 cases due to a lack of imaging findings text in the presented cases, and ultimately a total of 106 cases were included in this study. Each patient’s medical history and images (excluding pathological images) were collected from the “Question” section and the descriptions of imaging findings were collected from the “Answer” section of each published case. The “Answer” section contained descriptions of biopsy/surgical findings, histopathological findings, final/differential diagnoses, and discussion of diagnosis; thus, we excluded these descriptions from imaging findings. The data collection flowchart is presented in Fig. 1.

**Fig. 1.**
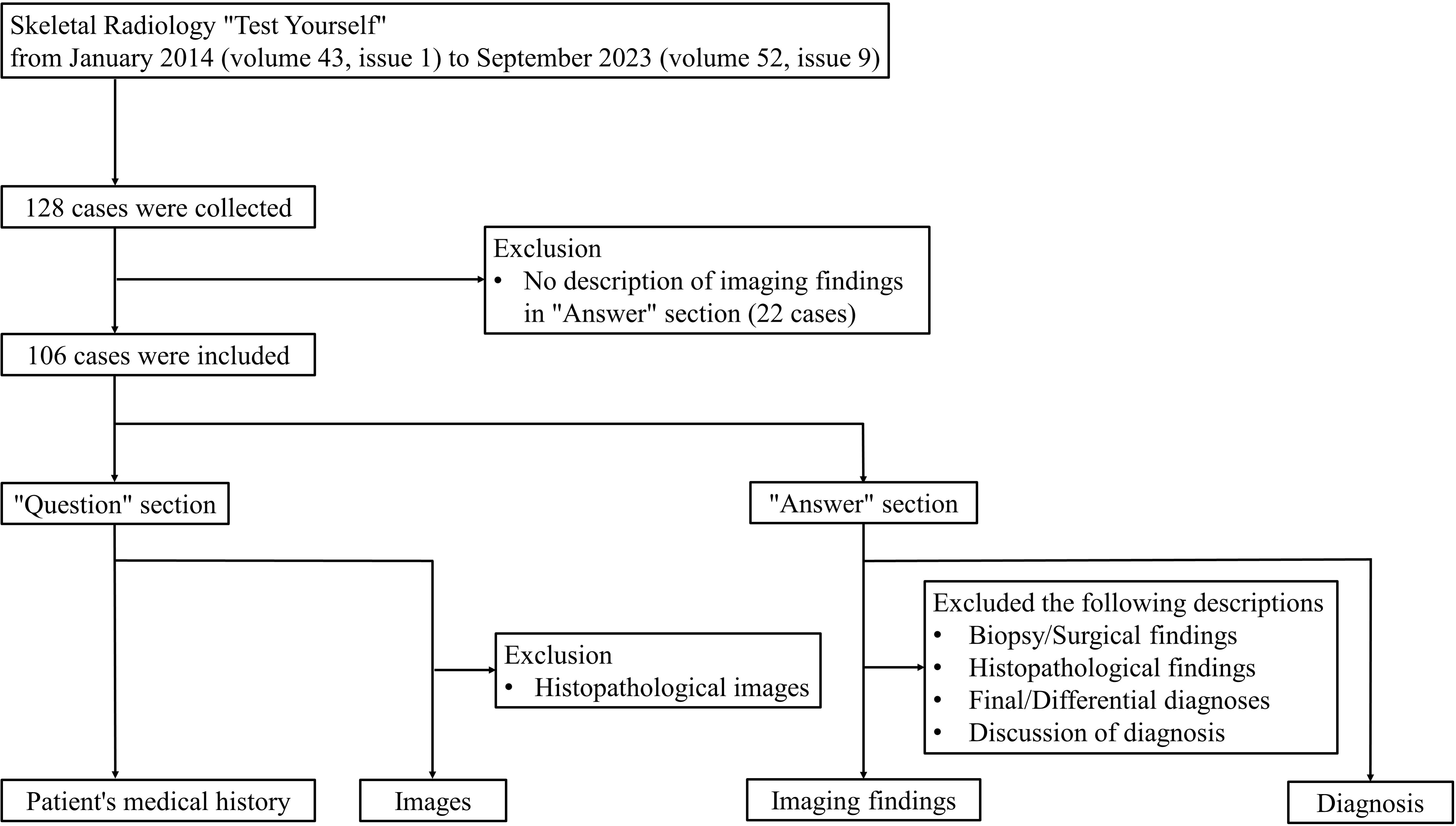
Data collection flowchart

### Input and output procedure for ChatGPT

First, the following premise was input into ChatGPT based on GPT-4 architecture (September 25 Version; OpenAI; https://chat.openai.com/) to prime it for the task: “As a physician, I plan to utilize you for research purposes. Assuming you are a hypothetical physician, please walk me through the process from differential diagnosis to the most likely disease step by step, based on the patient’s information I am about to present. Please list three possible differential diagnoses in order of likelihood” [14, 20]. Then, for GPT-4 based ChatGPT, the patient’s medical history and descriptions of imaging findings were input while, for GPT-4V based ChatGPT, the patient’s medical history and images themselves were input. The subsequent output from ChatGPT was collected (as shown in Fig. 2, 3). We started a new ChatGPT session for each case to prevent any potential influence of previous answers on ChatGPT’s output. These procedures were performed once for each case between September 28 and October 6, 2023.

**Fig. 2.**
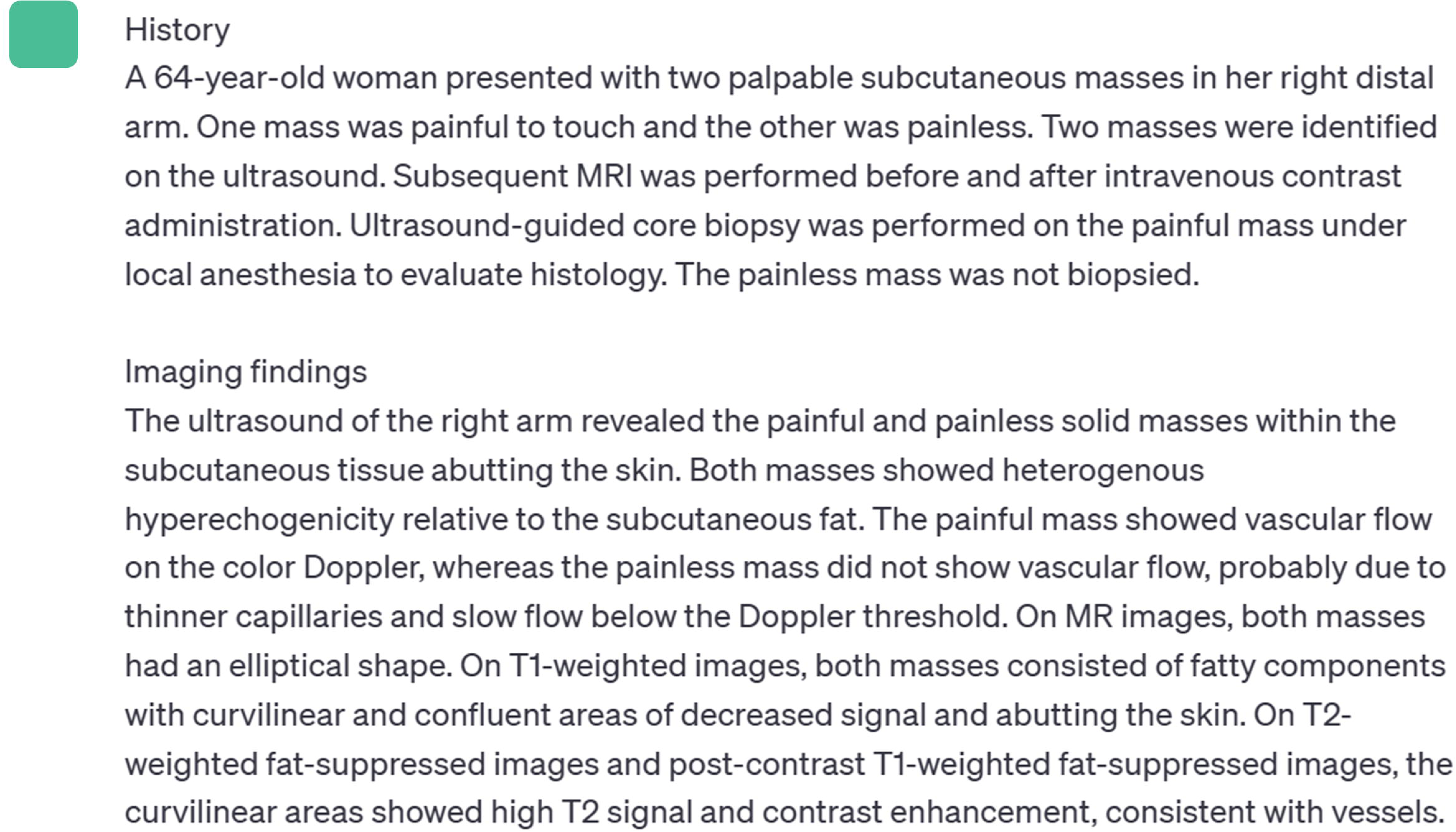
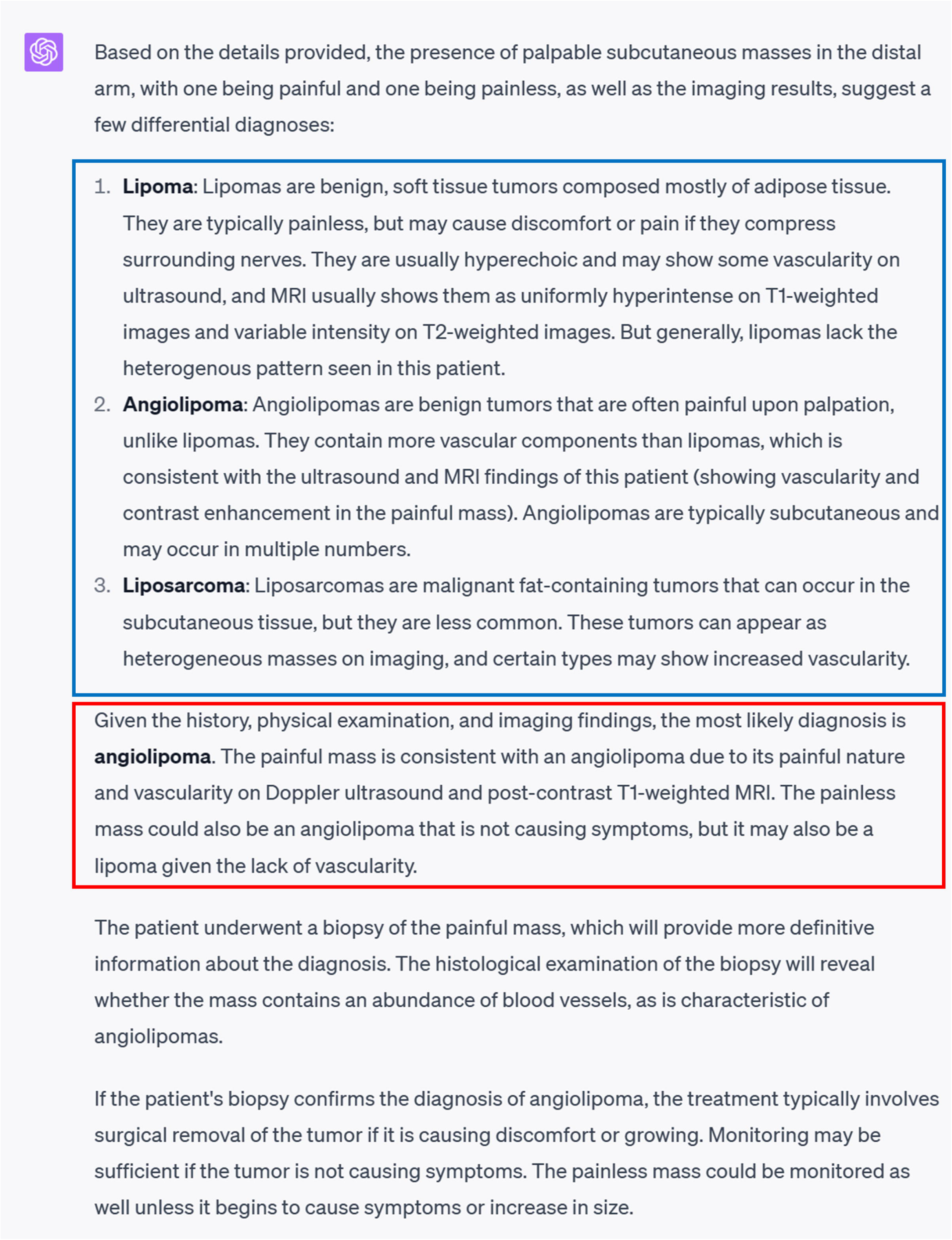
Input (patient’s medical history and imaging findings) and output examples of GPT-4 based ChatGPT. a Input texts to ChatGPT. b Output texts generated by ChatGPT. The differential diagnoses are outlined in blue and the final diagnosis is outlined in red. The final diagnosis generated by ChatGPT is correct in this case [31] [32].

**Fig. 3.**
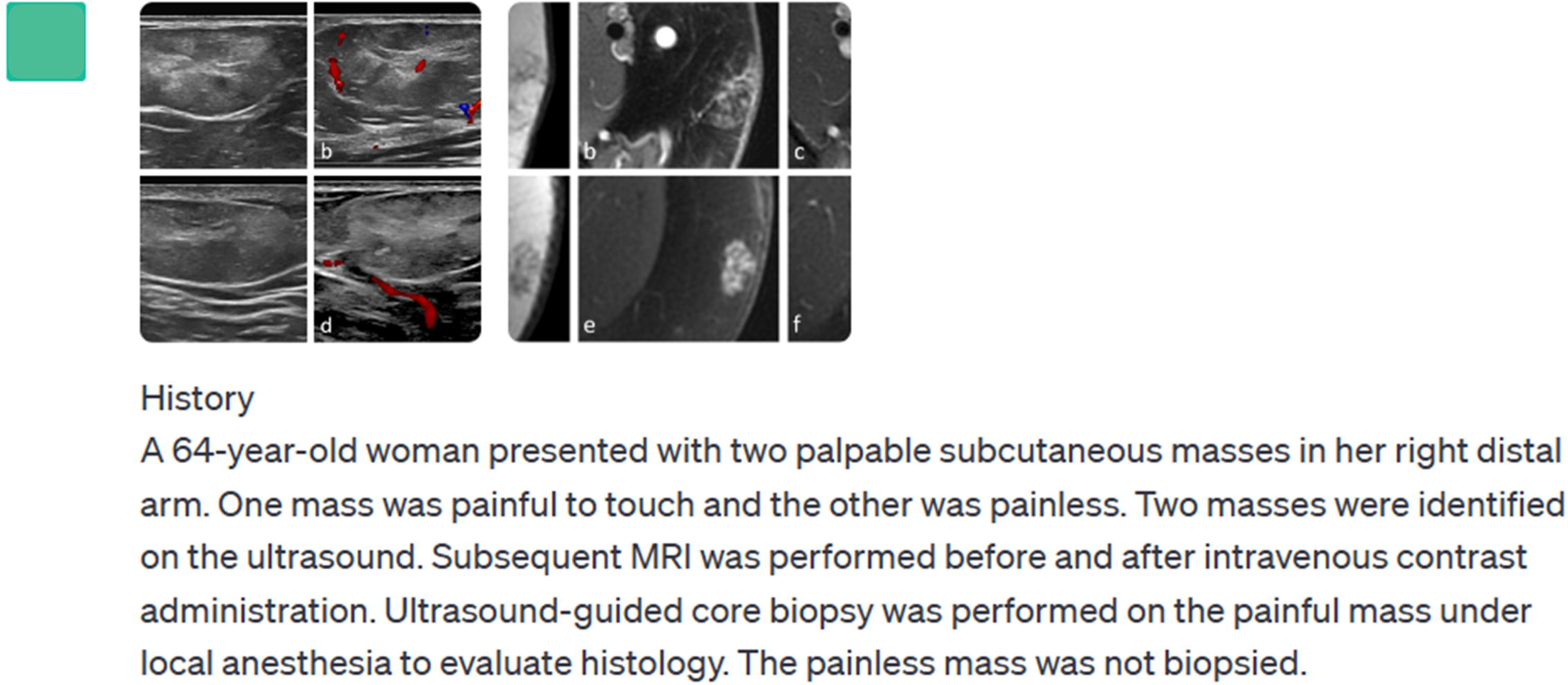
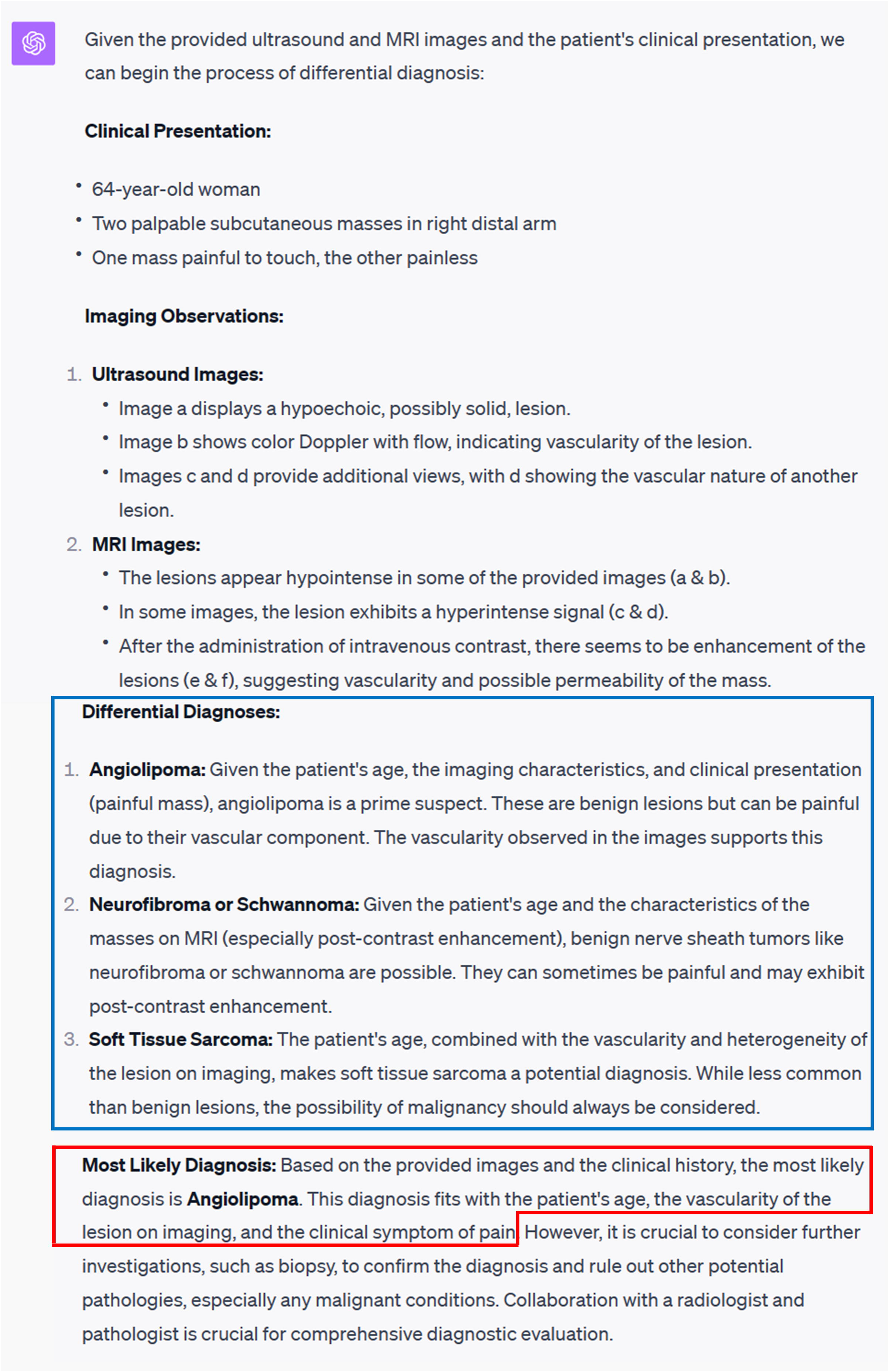
Input (patient’s medical history and images) and output examples of GPT-4V based ChatGPT. a Input to ChatGPT. b Output texts generated by ChatGPT. The differential diagnoses are outlined in blue and the final diagnosis is outlined in red. The final diagnosis generated by ChatGPT is correct in this case [31] [32].

### Output evaluation and category classification

The output generated by GPT-4 based ChatGPT and GPT-4V based ChatGPT included three differential diagnoses and one final diagnosis. Two board-certified radiologists (13 years of experience [H.T.]; 7 years of experience [D.H.]) evaluated both the differential diagnoses and the final diagnosis generated by ChatGPT to determine whether they were consistent with the actual ground truth in consensus. Each case was categorized into two groups: the tumor group and the non-tumor group, according to the 2020 World Health Organization classification of soft tissue and bone tumours [21]. The cases in the tumor group were further divided into bone tumor and soft tissue tumor cases.

### Radiologists’ interpretation

Two radiologists with different levels of experience (Reader 1 [T.O.]; a radiology resident with 4 years of experience) and (Reader 2 [D.H.]; a board-certified radiologist with 7 years of experience) independently reviewed all 106 cases. Both radiologists conducted their diagnoses based on the patient’s medical history and images (from the “Question” section). They provided three differential diagnoses and chose one as the final diagnosis for each case, and the diagnostic accuracy rates were evaluated. Both radiologists were blinded to the actual ground truth, as well as the differential and final diagnoses generated by ChatGPT.

### Statistical analysis

Statistical analyses were performed using R software (version 4.0.2, 2020; R Foundation for Statistical Computing; http://www.r-project.org/). Chi-square tests were conducted to compare the final and differential diagnostic accuracy rates between GPT-4 based ChatGPT and GPT-4V based ChatGPT. Chi-square tests were also conducted to compare the final and differential diagnostic accuracy rates between GPT-4 based ChatGPT and each radiologist, as well as between GPT-4V based ChatGPT and each radiologist. Furthermore, ChatGPT’s final and differential diagnostic accuracy rates for 1) the tumor and non-tumor groups, and 2) the bone tumor and soft tissue tumor cases were compared with pairwise Fisher’s exact tests. Adjustment for multiplicity was not performed because this was an exploratory study. *P* < 0.05 was considered statistically significant.

## Results

### ChatGPT’s diagnostic accuracy: GPT-4 based ChatGPT vs GPT-4V based ChatGPT

In all 106 cases, GPT-4 based ChatGPT (based on the patient’s medical history and imaging findings) and GPT-4V based ChatGPT (based on the patient’s medical history and images) successfully generated three differential diagnoses and provided one final diagnosis. GPT-4 based ChatGPT’s diagnostic accuracy rates for the final and differential diagnoses were 43% (46/106) and 58% (62/106), respectively. In contrast, GPT-4V based ChatGPT’s diagnostic accuracy rates for the final and differential diagnoses were 8% (9/106) and 14% (15/106), respectively. Both the final and differential diagnostic accuracy rates were significantly higher for GPT-4 based ChatGPT compared to GPT-4V based ChatGPT (*p* < 0.001 and < 0.001, respectively).

### Comparison of the diagnostic accuracy between ChatGPT and radiologists

Regarding the radiologists’ diagnostic accuracy, Reader 1 (a radiology resident) achieved a final diagnostic accuracy of 41% (43/106) and a differential diagnostic accuracy of 58% (61/106). Reader 2 (a board-certified radiologist) achieved a final diagnostic accuracy of 53% (56/106) and a differential diagnostic accuracy of 67% (71/106).

GPT-4 based ChatGPT’s diagnostic accuracy rates for the final and differential diagnoses were comparable to those of Reader 1 (*p* = 0.78 and 0.99, respectively), but lower than those of Reader 2, though not significantly (*p* = 0.22 and 0.26, respectively) (Table 1) (Fig. 4). In contrast, GPT-4V based ChatGPT’s diagnostic accuracy rates for the final and differential diagnoses were significantly lower than those of both radiologists (all *p* < 0.001).

**Fig. 4.**
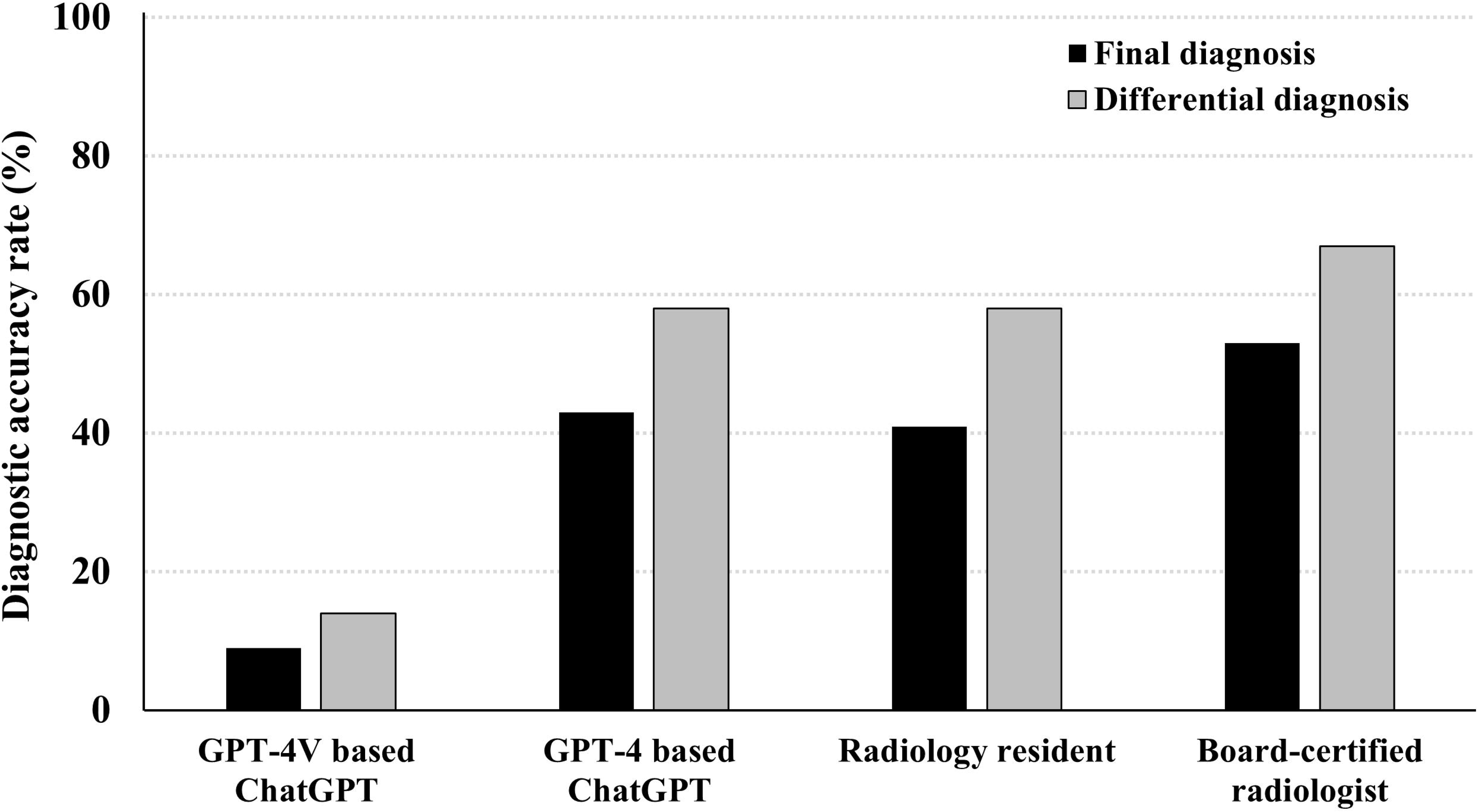
Diagnostic accuracy of GPT-4 based ChatGPT, GPT-4V based ChatGPT, and radiologists

**Table 1.** Comparison of the diagnostic accuracy between ChatGPT and radiologists.

|  | Correct answer (accuracy rate [%]) |  |  |  |
| --- | --- | --- | --- | --- |
|  | Final diagnosis | *p* value* | Differential diagnosis | *p* value* |
| GPT-4 based ChatGPT | 46/106 (43%) |  | 62/106 (58%) |  |
| Reader 1 (Radiology resident) | 43/106 (41%) | 0.78 | 61/106 (58%) | 0.99 |
| Reader 2 (Board-certified radiologist) | 56/106 (53%) | 0.22 | 71/106 (67%) | 0.26 |
| GPT-4V based ChatGPT | 9/106 (8%) |  | 15/106 (14%) |  |
| Reader 1 (Radiology resident) | 43/106 (41%) | < 0.001 | 61/106 (58%) | < 0.001 |
| Reader 2 (Board-certified radiologist) | 56/106 (53%) | < 0.001 | 71/106 (67%) | < 0.001 |

\* Chi-square tests are performed to compare the accuracy rates between GPT-4 based ChatGPT and each radiologist, as well as between GPT-4V based ChatGPT and each radiologist

### Categorical analysis of ChatGPT’s diagnostic accuracy

Detailed diagnostic accuracy rates for ChatGPT are shown in Table 2. Given the limited number of correct diagnoses by GPT-4V based ChatGPT, a categorical analysis was considered inappropriate due to the limited statistical power. Thus, we conducted a categorical analysis only for GPT-4 based ChatGPT’s diagnostic accuracy.

**Table 2.** ChatGPT’s diagnostic accuracy categorized by etiology.

|  | Correct answer (accuracy rate [%]) |  |  |  |
| --- | --- | --- | --- | --- |
|  | GPT-4 based ChatGPT |  | GPT-4V based ChatGPT |  |
|  | Final diagnosis | Differential diagnosis | Final diagnosis | Differential diagnosis |
| Total (n = 106) | 46/106 (43%) | 62/106 (58%) | 9/106 (8%) | 15/106 (14%) |
| Tumor group (n = 45) | 14/45 (31%) | 22/45 (49%) | 4/45 (9%) | 5/45 (11%) |
| Non-tumor group (n = 61) | 32/61 (52%) | 40/61 (66%) | 5/61 (8%) | 10/61 (16%) |
| Tumor group (n = 45)* | 14/45 (31%) | 22/45 (49%) | 4/45 (9%) | 5/45 (11%) |
| Bone tumor (n = 24) | 8/24 (33%) | 14/24 (58%) | 2/24 (8%) | 3/24 (13%) |
| Soft-tissue tumor (n = 22) | 6/22 (27%) | 9/22 (41%) | 2/22 (9%) | 2/22 (10%) |

\* One case presents both a bone tumor and a soft tissue tumor

When comparing the tumor and non-tumor groups, the final and differential diagnostic accuracy rates were 31% (14/45) and 49% (22/45) for the tumor group, and 52% (32/61) and 66% (40/61) for the non-tumor group, respectively. The tumor group showed significantly lower final diagnostic accuracy rates compared to the non-tumor group (*p* = 0.03), while there was no significant difference between the differential diagnostic accuracy rates of the two groups (*p* = 0.11). Within the tumor group, the final and differential diagnostic accuracy rates were 33% (8/24) and 58% (14/24) in bone tumor cases, and 27% (6/22) and 41% (9/22) in soft tissue tumor cases, respectively (one presented both a bone tumor and a soft tissue tumor). When comparing the diagnostic accuracy rates between bone tumor and soft tissue tumor cases, no significant difference was observed in either the final or differential diagnosis (*p* = 0.75 and 0.38, respectively).

## Discussion

This study demonstrated the diagnostic accuracy of GPT-4 based ChatGPT and GPT-4V based ChatGPT in musculoskeletal radiology. The diagnostic accuracy of GPT-4 based ChatGPT (based on the patient’s medical history and imaging findings) was significantly higher than that of GPT-4V based ChatGPT (based on the patient’s medical history and images). Regarding the comparison between ChatGPT and radiologists, GPT-4 based ChatGPT’s diagnostic accuracy was comparable to that of a radiology resident but lower than that of a board-certified radiologist. While GPT-4V based ChatGPT’s diagnostic accuracy was significantly lower than that of both radiologists. In the analysis of GPT-4 based ChatGPT’s diagnostic accuracy per category, GPT-4 based ChatGPT’s final diagnostic accuracy rate was significantly lower for the tumor group compared to the non-tumor group. Within the tumor group, the accuracy rates for the final and differential diagnoses were relatively higher for bone tumor cases compared to those of soft tissue tumor cases, although the differences were not significant.

To the best of our knowledge, this study is the first in the field of musculoskeletal radiology to investigate the diagnostic capability of GPT-4 and GPT-4V based ChatGPTs and to compare these to radiologists’ performance. Although a previous study has reported that GPT-3 based ChatGPT can generate coherent research articles in musculoskeletal radiology [18], no study has evaluated the diagnostic performance of GPT-4 and GPT-4V based ChatGPTs in this field. This study provides valuable insights into the strengths and limitations of using ChatGPT as a diagnostic tool in musculoskeletal radiology.

ChatGPT has the potential to enhance diagnostic accuracy and consequently improve the diagnostic workflow in musculoskeletal radiology [22]. The exponential growth of medical imaging technologies and the overutilization of imaging examinations have substantially increased musculoskeletal imaging, subsequently increasing the workload for radiologists [23, 24]. The higher workload for radiologists not only leads to diagnostic errors but also affects job satisfaction, contributing to burnout [25, 26]. The implementation of ChatGPT as a diagnostic support tool has the potential to optimize the diagnostic imaging process, resulting in time savings and a decreased workload for radiologists, thereby increasing overall efficiency, reducing diagnostic errors, and ultimately improving patient outcomes. In addition, ChatGPT could be valuable in a clinical setting where expertise in musculoskeletal radiology is limited, as it is easily accessible at any time and from anywhere.

While ChatGPT holds promise for revitalizing musculoskeletal radiology, radiologists should recognize its capabilities and exercise caution when incorporating ChatGPT into clinical practice. This study demonstrated that the diagnostic accuracy of GPT-4 based ChatGPT was significantly higher than that of GPT-4V based ChatGPT. These results indicated that the GPT-4V based ChatGPT’s capability to process images and extract imaging findings is insufficient. In OpenAI’s statements, they considered the current GPT-4V to be unsuitable for performing the interpretation of medical images and replacing professional medical diagnoses due to inconsistencies [5]. Therefore, it is essential to input appropriate descriptions of imaging findings when using ChatGPT as a diagnostic tool in clinical practice. Regarding the comparison between ChatGPT and radiologists, GPT-4V based ChatGPT’s diagnostic performance was significantly lower than that of radiologists, and GPT-4 based ChatGPT’s diagnostic performance was comparable to that of radiology residents but did not reach the performance level of board-certified radiologists. Although ChatGPT may assist radiologists in narrowing down differential diagnoses, ChatGPT alone cannot fully replace the expertise of radiologists and should only be used as an adjunct tool.

This study also revealed that the diagnostic accuracy of GPT-4 based ChatGPT may vary depending on the etiology of the disease; it was significantly lower in the tumor group compared to the non-tumor group. This lower diagnostic accuracy in neoplastic diseases could be attributed to the challenging nature of interpreting complex cases, due to the wide variety of histopathological types and imaging findings [21, 27]. Rare neoplastic diseases may be more challenging for ChatGPT due to the limited literature and a lack of established typical imaging findings. Although no significant difference in diagnostic accuracy rates was observed between bone tumor and soft tissue tumor cases, bone tumor cases showed relatively higher accuracy rates compared to soft tissue tumor cases. While soft tissue tumors of both benign and malignant nature often share overlapping imaging features [28], bone tumors have grading systems that allow for the assessment of malignancy risk based on their growth patterns [29, 30]. This distinction may be one of the contributing factors to the relatively higher differential diagnostic accuracy for bone tumors compared to soft tissue tumors. On the other hand, the significantly higher accuracy rates for the final diagnosis of the non-tumor group indicated that GPT-4 based ChatGPT may be particularly useful in diagnosing non-neoplastic diseases in musculoskeletal radiology.

This study had several limitations. First, ChatGPT’s performance in generating diagnoses was conducted in the controlled environment of the “Test Yourself” cases, which may not fully represent the broader range of musculoskeletal radiology cases. This selection bias could affect the generalizability of the results and may not capture the full spectrum of diagnostic challenges encountered in real-world clinical practice. Second, the “Test Yourself” cases represent a potential for bias since these cases may have been included in the training data of ChatGPT. This bias may lead to an overestimation of ChatGPT’s diagnostic accuracy. Third, this study did not conduct a categorical analysis for GPT-4V based ChatGPT’s diagnostic accuracy due to the limited number of correct diagnoses which limits the statistical power of the analyses. Fourth, this study did not evaluate ChatGPT’s diagnostic accuracy by further subdividing etiologies in non-neoplastic diseases due to the limited number of cases.

In conclusion, this study demonstrated the diagnostic accuracy of not only GPT-4 based ChatGPT but also GPT-4V based ChatGPT in musculoskeletal radiology. The diagnostic accuracy of GPT-4 based ChatGPT was significantly higher than that of GPT-4V based ChatGPT; therefore, it is essential to input appropriate descriptions of imaging findings when using ChatGPT as a diagnostic tool in clinical practice. The diagnostic performance of GPT-4 based ChatGPT was comparable to that of radiology residents but did not reach the performance level of board-certified radiologists. While ChatGPT may assist radiologists in narrowing down the differential diagnosis and improving the diagnostic workflow, radiologists need to be aware of its capabilities and limitations for optimal utilization.

## Data Availability

All data produced in the present work are contained in the manuscript

## Acknowledgments

Our manuscript was developed with the assistance of ChatGPT, a language model based on the GPT-4 architecture (September 25 Version; OpenAI; https://chat.openai.com/). However, all outputs generated by ChatGPT were reviewed and approved by the authors.

## Compliance with Ethical Standards

### Funding

This study was supported by Guerbet.

### Ethical approval

All procedures performed in studies involving human participants were in accordance with the ethical standards of the institutional and/or national research committee and with the 1964 Helsinki declaration and its later amendments or comparable ethical standards. This study was approved by the institutional review board of our institution, and informed consent was not required since this study utilized only published cases.

